# Optimal allocation of PCR tests to minimise disease transmission through contact tracing and quarantine

**DOI:** 10.1101/2021.03.23.21254148

**Authors:** Christopher M. Baker, Iadine Chades, Jodie McVernon, Andrew Robinson, Howard Bondell

## Abstract

PCR testing is a crucial capability for managing disease outbreaks, but it is also a limited resource and must be used carefully to ensure the information gain from testing is valuable. Testing has two broad uses, namely to track epidemic dynamics and to reduce transmission by identifying and managing cases. In this work we develop a modelling framework to examine the effects of test allocation in an epidemic, with a focus on using testing to minimise transmission. Using the COVID-19 pandemic as an example, we examine how the number of tests conducted per day relates to reduction in disease transmission, in the context of logistical constraints on the testing system. We show that if daily testing is above the routine capacity of a testing system, which can cause delays, then those delays can undermine efforts to reduce transmission through contact tracing and quarantine. This work highlights that the two goals of aiming to reduce transmission and aiming to identify all cases are different, and it is possible that focusing on one may undermine achieving the other. To develop an effective strategy, the goals must be clear and performance metrics must match the goals of the testing strategy. If metrics do not match the objectives of the strategy, then those metrics may incentivise actions that undermine achieving the objectives.

## Introduction

Testing is an important aspect of monitoring and managing an epidemic, because it provides information to set policy and allows us to reduce transmission by managing cases (Salathé et al. 2020). Tests are the primary way of tracking pandemic progress, and the data are used to fit dynamical models and estimate the rate of community spread (Abbott et al. 2020; Moss et al. 2020). Understanding the dynamics of the spread allows governments to be better informed when setting policy and preparing healthcare capacity. Alongside the policy implications of testing, the tests themselves can reduce transmission. If infectious individuals are identified as cases, then they can be isolated and their close contacts quarantined to limit onward spread of infection (Kretzschmar et al. 2020; Larremore et al. 2020; Mina et al. 2020).

Testing policies and aims vary globally, although there is a common focus on surveillance to both track the epidemic and to reduce spread. These two general aims are often split into more specific objectives, for example, the European Centre for Disease Prevention and Control (ECDC) define 5 objectives: (i) control transmission; (ii) monitor incidence and trends and assess severity over time; (iii) mitigate the impact of COVID-19 in healthcare and social-care settings; (iv) rapidly identify all clusters or outbreaks in specific settings; and (v) prevent (re-)introduction into regions/countries with sustained control of the virus (European Centre for Disease Prevention and Control 2020). The EDCD document says that to control transmission effectively, a test should have an action linked to the result, and test results should be known within 24 hours to ensure timely action (European Centre for Disease Prevention and Control 2020).

In testing policy documents, there appears to be little discussion about system capacity and how testing strategy should be tailored depending on capacity and system lags. The omission of capacity and the related delays is a clear gap, because we know timeliness is critical and contact tracing works best when the system is working efficiently with minimal delays (Gardner & Kilpatrick 2020; Quilty et al. 2020). Rather, the advice is that capacity should be expanded such that all people with symptoms can be tested and additional testing can occur if there is remaining capacity (European Centre for Disease Prevention and Control 2020).

Despite the clear benefits of widespread testing, there are important trade-offs to consider when developing testing strategy. PCR testing is the global ‘gold standard’ for identifying cases (Shen et al. 2020), but it suffers from logistical challenges. Typically, a nasopharyngeal swab is taken by a healthcare provider, and the swab is sent for laboratory testing (Pondaven-Letourmy et al. 2020). There are various delays in the testing process, which we group into two categories, namely the *swab delay* and the *turnaround time (TAT)*. The swab delay is the time elapsed between a person developing symptoms and presenting for a test. The turnaround time (TAT) is the time between swab collection and the results being reported, which is when contacts of cases are typically managed.

The effectiveness of a testing strategy for both isolation and for quarantine of contacts depends on its timeliness. The earlier a person is identified as a case, relative to onset of their infectious period, the greater the reduction in transmission (Kretzschmar et al. 2020; Plank et al. 2020). The speed of the test result (TAT) should not influence the effectiveness of isolation in theory, as the person should isolate while awaiting results, despite its inconvenience, although there are cases of noncompliance (Smith et al. 2020). However, the effect of the TAT is most pronounced for contact tracing, as contacts are typically not quarantined until the case is confirmed.

Although PCR tests can be completed in under 24 hours (Ramdas et al. 2020), laboratories process only a limited number of tests per day, and testing beyond capacity leads to delays and increased TATs. For example, in New York there have been reports of TATs of greater than 6 days (Rosa 2020), and data from the UK shows increasing TATs when more tests are completed (see Supplementary Information S1). The effectiveness of quarantine depends on how quickly contact tracing occurs and whether infected individuals can be quarantined before they become infectious which, for COVID-19, precedes symptom onset (Kucharski et al. 2020). TATs that stretch into multiple days reduce the effectiveness of isolation and quarantine to limit transmission.

While there have been a range of analyses exploring testing strategy (Kretzschmar et al. 2020; Larremore et al. 2020), but these do not jointly incorporate testing delays and system capacity. Some early papers argued for fast and frequent testing as a strategy to reduce spread, even if test sensitivity is relatively low (Larremore et al. 2020; Mina et al. 2020), while other work noted that timeliness is critical, and contact tracing works best when the system is working efficiently (Gardner & Kilpatrick 2020). Kretzschmar et al. (2020) used a branching process model to quantify the achievable reduction in transmission from isolation and quarantine given variable swab delays and TATs, but did not explicitly link these lag times to test demand and health system capacity. Other analyses seek to find optimal testing strategies when the number of tests per day has a fixed upper limit (but no delays), which show that a good testing strategy can both deliver better information about the epidemic dynamics (Chatzimanolakis et al. 2020) and reduce disease spread (Jonnerby et al. 2020).

In this paper we develop a framework for optimising test strategy when seeking to minimise community transmission. We incorporate testing delays into our model, which depend on test demand and lab capacity. Using testing delays, rather than setting a fixed upper limit on testing, means that we get an explicit trade-off between testing volume and speed. Using previous results about the impact of testing on transmission (Kretzschmar et al. 2020), we identify the optimal number of tests per day, depending on community prevalence and test system capacity. In doing so, we show there is a delicate balance to strike when optimising use of limited public health resources and that more testing is not always better.

## Methods

Our motivating question is: on a given day, how should tests be distributed across the population to minimise future transmission from currently-unidentified infections? We answer this question by quantifying the *value of a test* for a person, in terms of reduced onward transmission of infection, and then describe how to allocate tests to the population to maximise the overall value of the strategy. We break the problem into 3 parts:

1. Quantifying the TAT, as a function of test demand and lab capacity;
2. Relating TAT to reduction in onwards infection transmission; and
3. Optimally allocating tests to minimise future transmission.

### Test volume and turnaround time (TAT)

The link between the number of daily tests and the TAT is fundamental to this work. We use a simple model to represent the testing system, in which we assume there is a routine capacity and a corresponding baseline TAT. Whenever the number of tests per day is under routine capacity, the time until results is equal to the baseline TAT, which we generally set to be 1 day. This timeframe corresponds to what NSW Health, Australia has achieved in 2020 for COVID-19 (NSW Health 2020). When testing goes beyond routine capacity, into *surge capacity*, we assume the TAT increases. While there is evidence that TATs increase with increased test volume from the UK (see Supplementary Information S1), the precise relationship is unclear and may vary between jurisdictions. Hence, we use a quadratic relationship between TAT and excess tests in the main figures of this work (Figure 1), but we also replicate our work with linear and exponential relationships to show that our qualitative conclusions are robust to the function choice (see Supplementary Information S2).

**Figure 1.**
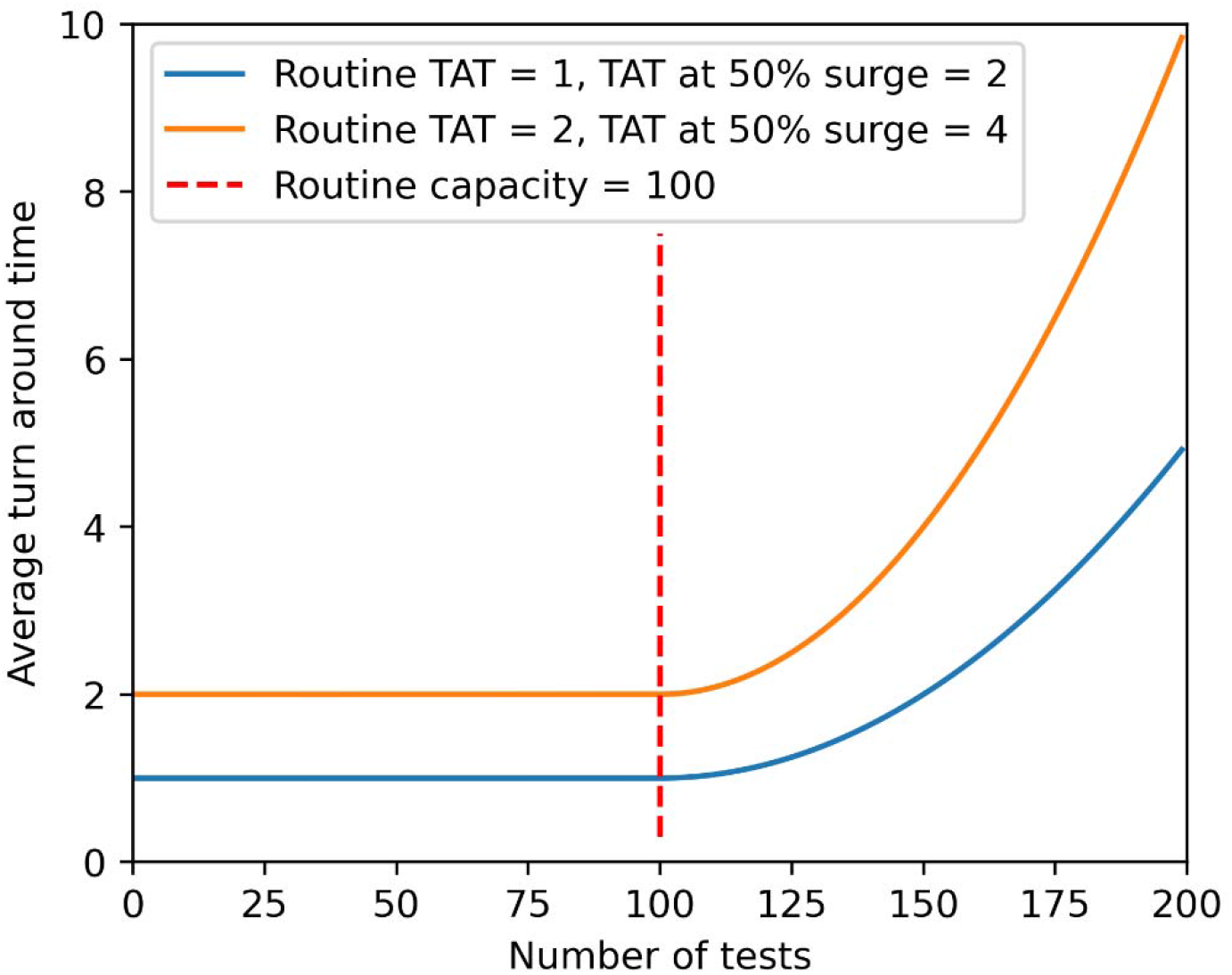
Modelled relationship between the number of tests (daily) and the turnaround time (TAT), with routine capacity set at 100 tests per day (red dashed line). Throughout this work, we use a routine TAT of 1 days and a TAT at 50% surge capacity of 2 days (solid blue line) unless otherwise stated. The orange line shows another example of the type of relationship between test volume and TAT that this model allows.

#### Turnaround time and transmission reduction

Understanding the link between TAT and transmission is vital for determining the optimal testing strategy. We use the model developed by Kretzschmar et al. (2020) to quantify the percentage reduction in transmission for a person, as a function of swab delay and TAT (Figure 2). We consider two components to the transmission reduction, namely: the impact of case isolation, and the impact of contact tracing via quarantine of contacts. The swab delay has a big impact on overall transmission and, if the swab delay is small, then isolation alone can reduce transmission greatly. Quarantine of contacts gives a further reduction in transmission and again the best results are realised when the TAT is very low. If the TAT were 7 days or more, quarantine of contacts would have almost no additional benefit, so the transmission reduction arises only from isolation of the case.

**Figure 2.**
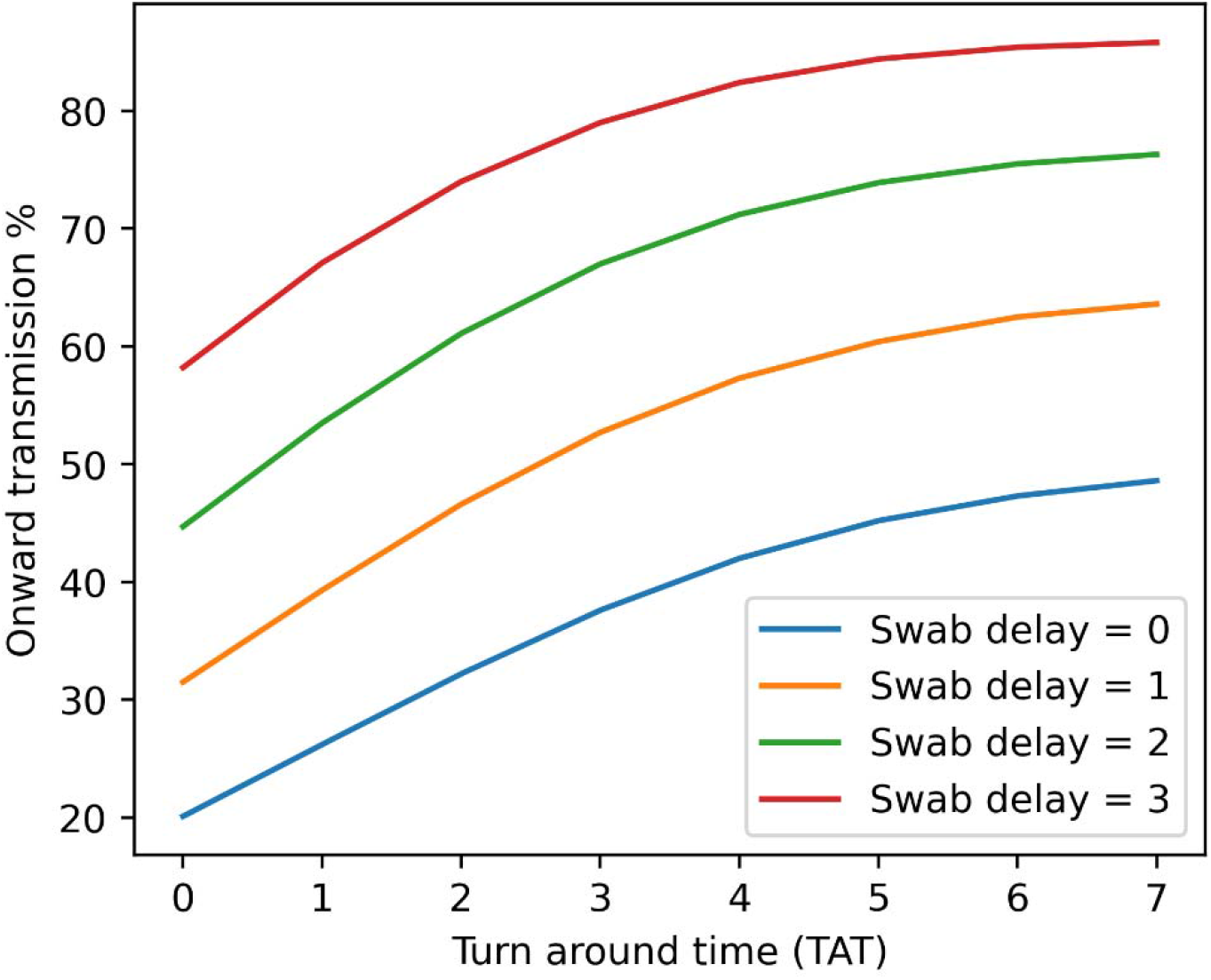
The reduction in onwards transmission through testing, using the model from Kretzschmar et al. (2020), under the assumption that a person isolates while awaiting the test result. We show the reduction in onwards transmission for varying swab delays, from 0 days at the bottom (blue line) to 3 days at the top (red line). Although the absolute reduction in transmission differs greatly depending on the swab delay, the difference between a TAT of 0 days and 7 days is approximately 25% for this range of swab delays. Note: we extended the results presented in Kretzschmar et al. (2020) Table 2 by re-running their Mathematica code, increasing the maximum value of to 7, and interpolating the curve in between the integer days.

**Table 1:** The number of close contacts, symptomatics and asymptomatics for the outbreak response and community transmission scenarios

| **Number of people per 100,000** | *Close contact* | *Symptomatic* | *Asymptomatic* |
| --- | --- | --- | --- |
| *Outbreak response* | 14 | 600 | 99,386 |
| *Community transmission* | 140 | 600 | 99,260 |

**Table 2:** The probability of testing positive for close contacts, symptomatics and asymptomatics for the outbreak response and community transmission scenarios.

| **Probability of positive test** | *Close contact* | *Symptomatic* | *Asymptomatic* |
| --- | --- | --- | --- |
| *Outbreak response* | 2% | 0.05% | 0.0001% |
| *Community transmission* | 2% | 0.5% | 0.001% |

### Test allocation and indications

We next identify the optimal number of tests to conduct, alongside who should be tested. We stratify the population into three groups or *indications*: close contacts, symptomatics and asymptomatics. Close contacts are defined as people who have been in contact with a case identified on a previous day, symptomatics are people who are showing symptoms consistent with the disease, and asymptomatics are people who are not showing symptoms. Each of these indications will have a different probability of testing positive and different onwards transmission rates (for example, an infected close contact should have lower onwards transmission rate than an infected symptomatic person, as known close contacts should already be quarantined).

For each person, we define *R_E_*,, in the absence of a test, given the person has the disease. We define *R_T_* as the expected onwards transmission with a test:

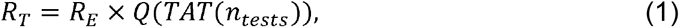

where *Q* is the proportion of the expected onwards transmission, given a test (Figure 2), which is a function of the TAT, and *TAT*(*n_tests_*) is the TAT when the number of tests done per day is *n_tests_* (e.g. Figure 1).

To estimate the expected value of the test, we must also include the likelihood that the person has the disease, denoted P. The value of a test is:

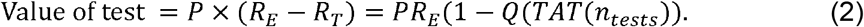

The likelihood of being infected varies between groups of people, and therefore the value of a test also varies, depending on the person. For a set number of tests, N, per day, we then estimate the value of a test for each group in the population and order the population groups from most valuable to least valuable to test, and test those that are most valuable.

Based on this testing protocol, by varying the number of daily tests, *n_tests_*, the percentage of positive tests and overall transmission reduction can be calculated for each possible *n_tests_*. This enables a comparison across the number of tests in order to identify the optimal number of tests per day, which is done by choosing *n_tests_* to minimise Eq. (1), and prioritising those tests using Eq. (2).

## Results

The testing strategy depends on the logistics of the testing system, the prevalence of disease in a population and the overall community restrictions, which affect the average onwards transmission without a test. We present results for two prevalence scenarios, which we term *outbreak response* and *community transmission* and are inspired by outbreaks in Australia, 2020. We also only focus on two levels of test capacity and, unless stated, assume that only 50% of symptomatic people will volunteer for testing (FluTracking 2020) (we also test the robustness of our conclusions to 25% and 75%, see Supplementary Information S3).

We report results for each prevalence scenario for two indicative testing systems. We consider two values for routine testing capacity: 2 tests per 1000 of population and 4 tests per 1000 of population, which are motivated by the test numbers reported in Australia (Macali 2021). For all of our results, we use a swab delay (time from symptom onset to swab collection) of 1 day, a baseline TAT of 1 day, and a surge TAT of 2 days when demand is 150% of capacity.

We show results for our two prevalence scenarios of *outbreak response* and *community transmission*. Our parameter choices were motivated by the epidemic in Australia, where cases peaked at approximately 14 per 100,000 with 2.8% positive test results in Victoria on the 5^th^ of August 2020 (Macali 2021). In the outbreak response setting, there are approximately 0.7 cases per 100,000 per day, while in the community transmission setting there are approximately 6.8 cases per 100,000 per day, and the number of close contacts in the population, the probability of returning a positive test and the average onward transmission without a test varies between the settings, as shown in Tables 1, 2 and 3. Our choices for parameter values are illustrative and we include additional figures to show the sensitivity of our results to each of onwards transmission, the number of people in each group and the pre-test probability (see Supplementary Information S4). We chose the number of close contacts to be approximately 20 per infection, and we set the number of symptomatics to be 600, which corresponds to an average of two illnesses per year per person. We assumed that the probability of a close contact testing positive is 2%, and that close contacts are the most likely indication to test positive. For symptomatics and asymptomatics we assumed that the probability of testing positive is an order of magnitude higher in when there is community transmission, compared to outbreak response. Finally, we set the baseline average number of onwards transmissions, for a person who doesn’t get identified as a case. We chose values for onwards transmission to be higher in outbreak response compared to community transmission, which assumes that when prevalence is higher, there are more restrictions to reduce transmission. We also assume that close contacts have reduced onwards transmission as they should be in quarantine. Our results are presented in terms of percentage reduction in transmission, so it is the relative values of average onward transmission that are important, rather than their absolute values.

**Table 3:** The average onward transmission for close contacts, symptomatics and asymptomatics for the outbreak response and community transmission scenarios.

| **Average onward transmission without a test** | *Close contact* | *Symptomatic* | *Asymptomatic* |
| --- | --- | --- | --- |
| *Outbreak response* | 0.75 | 1.5 | 1.5 |
| *Community transmission* | 0.25 | 1.25 | 1.25 |

We found that the amount of onwards transmission depended on the number of tests, and there was an intermediate value of testing that minimised transmission (Figure 3). For the scenario with routine capacity of 2 tests per 1000 of population, our model suggested testing slightly above routine testing capacity, as is worth finding more cases, despite the small increase in the TAT. With our parameter values, the maximum number of symptomatic people that can be tested is 3 per 1000. With routine capacity at 4 tests per 1000, it was not optimal to test above routine capacity, as all close contacts and symptomatic people could be tested within capacity, and testing asymptomatic people returned very few cases, but would delay results for everyone.

**Figure 3.**
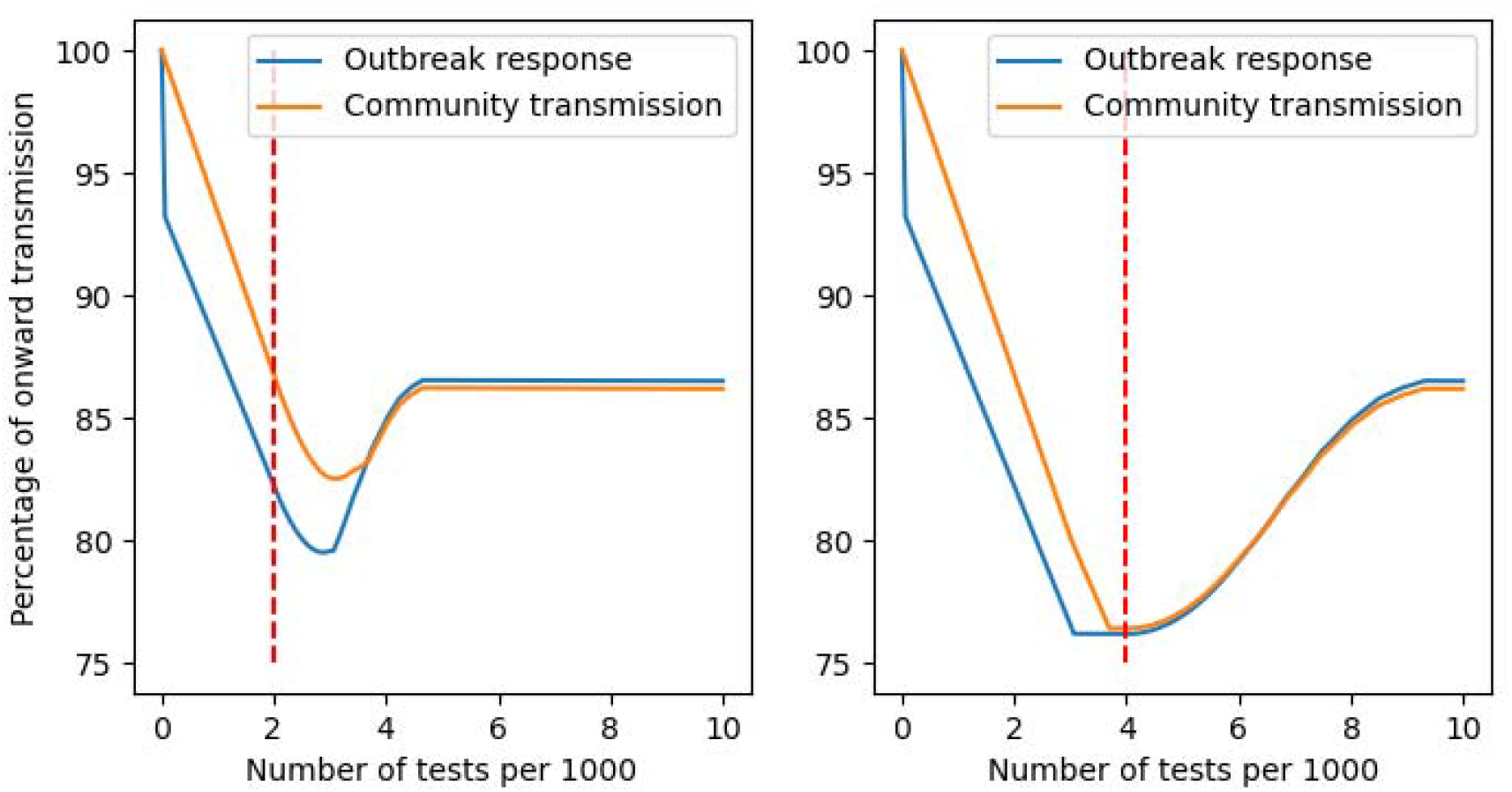
The percentage of transmission that occurs, depending on the number of tests done on a given day. The solid blue and orange lines represent the outbreak response and community transmission scenarios respectively. The left panel shows the results for the community transmission scenario with the test capacity set at 2 tests per 1000 people, while the right is for a test capacity of 4 tests per 1000 people.

The benefit of contact tracing and quarantine of close contacts was greatest when testing was within routine capacity (Figure 4). Within routine capacity, contact tracing provided approximately 40% of the benefits of testing, which decreased after routine capacity was reached. The decline in contact tracing effectiveness was initially small, but once test volume was more than double routine capacity, the effect of contact tracing was essentially zero. This result assumes that there are not follow-on constraints in contact tracing that further delay quarantine of close contacts, and any further delays would cause larger reductions in contact tracing effectiveness.

**Figure 4.**
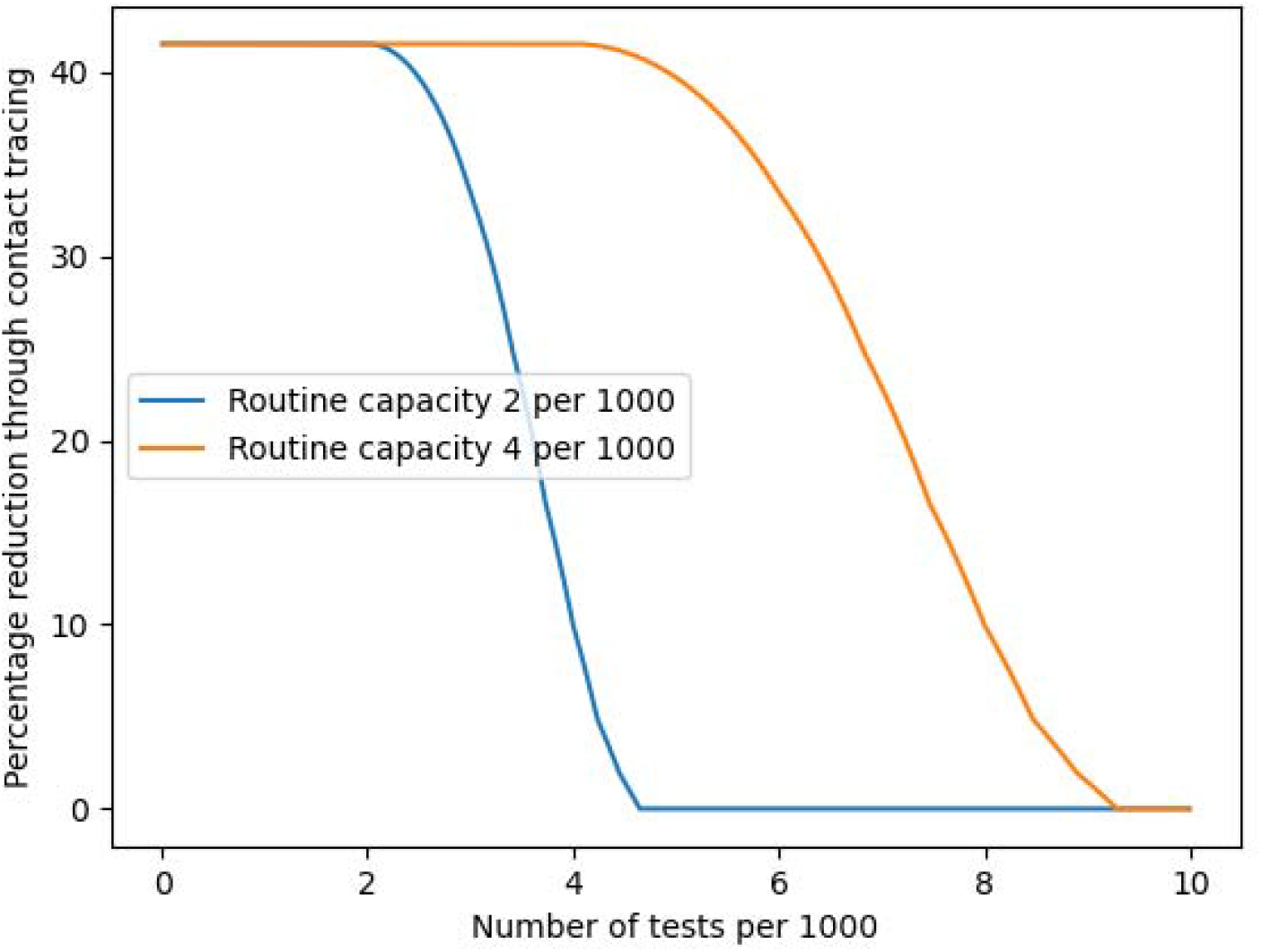
The percentage of the transmission reduction that was due to contact tracing and quarantine of contacts, the remaining being made up by the benefit of isolating the identified case. The benefit was greatest when test volume is below routine capacity and diminishes as daily tests increase into surge capacity. The plots for the outbreak response and community transmission were identical.

The percentage of tests that return positive results is not a good indicator for whether a testing strategy is effectively reducing transmission (Figure 5). For the community transmission scenario and routine capacity of 2 tests per 1000 of population, the optimal number of tests returns approximate 0.5% positive results, as only symptomatic people are tested. Increasing the testing would also increase the percent positive, as the second priority is close contacts, but would decrease if many asymptomatic people were tested. If the goals of a testing strategy include minimising the percentage positive tests, then there is an incentive to test widely, which could perversely affect transmission reduction.

**Figure 5.**
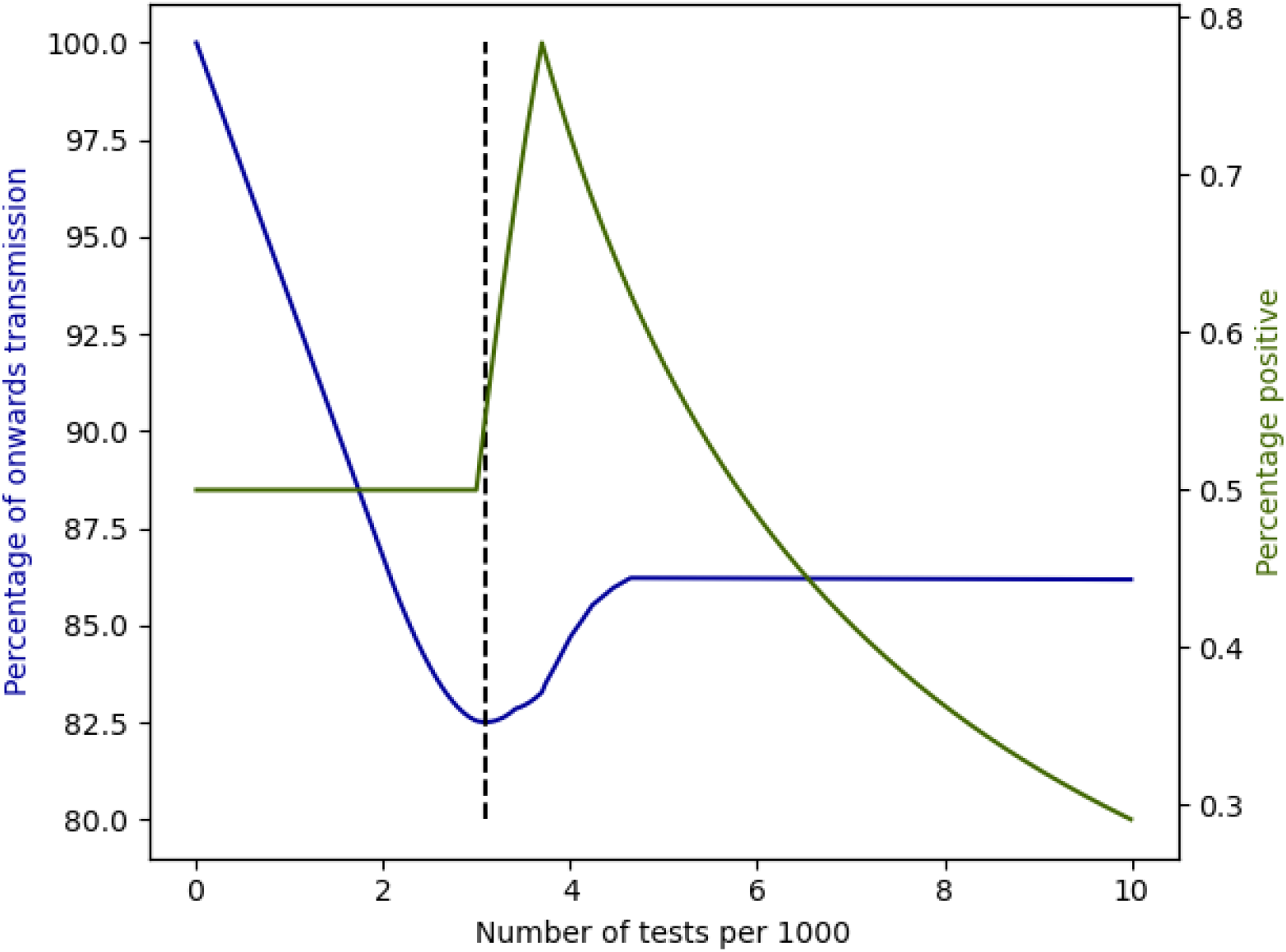
The percentage of onwards transmission (blue) and the percentage of tests that return positive results (dark green) for the community transmission scenario and routine capacity of 2 tests per 1000 per day. The vertical dashed black line shows the number of tests that minimises onwards transmission within these system constraints.

## Discussion

This work illustrates the balance required when determining a COVID-19 testing strategy. Conducting many tests is the only way to properly understand disease prevalence and identify changing dynamics, but unless the efficiency of the testing system can be improved to accommodate workload, too many tests slow down the system, inhibiting its value to reduce disease spread. We show that there is an optimum volume of testing if the goal is simply transmission reduction. That optimum depends on disease prevalence and on the testing system capacity. The key points are that test results must be fast enough to enable effective quarantine of close contacts of cases, highlighting the importance of the TAT as a key indicator of public health response efficacy. Focusing testing on the most valuable cohorts of a community (namely, those who are (1) likely to be infected and (2) have many close contacts) is the most effective way to reduce transmission. A strategy that aims to minimise transmission can mean that many infections are not identified as cases, if there is not sufficient system capacity to identify them and aiming to identify all cases could severely hamper control efforts.

Our work shows that an effective testing strategy can make a significant difference to disease transmission, even when there are realistic system capacities and delays. For an epidemic with a basic reproduction number (*R*_0_) of 2.5 – similar to early estimates for COVID-19 (Moss et al. 2020) – a 25% reduction in transmission (Figure 3) would reduce *R*_0_ to approximately 1.9. While this is a significant reduction, the testing system needs to work in unison with other transmission reduction policies to bring *R*_0_ below 1, or the testing system would need to be more effective that the one modelled here. The model also shows that an efficient testing system greatly reduces the number of tests required to attain a certain level of transmission reduction. For example, in the outbreak response scenario with routine capacity of 2 tests per 1,000 (Figure 3), the same outcome can be achieved with 2 tests per 1,000 per day as with 4 tests per 1,000 per day. The reason that testing 2 per 1,000 population achieves the same outcome is that the contact tracing is quick. An efficient system is not only useful because fewer people are required to isolate while awaiting results, it also reduces costs and reduces demand on the reagent supply.

This work examines how overloading the testing system affects our ability to control transmission, but testing systems are more intricate than our model. Our results show that the issue with too much testing is a less effective contact tracing and quarantine program (Figure 4). However, there may be strategies that involve within-system prioritisation of tests and contact tracing that could preserve contact tracing efficiency, while maintaining high testing volume to give people certainty and allow better tracking of epidemic progression. For example, there could be priority testing laboratories to ensure a proportion of tests were completed quickly and contact tracers could prioritise recent test results, to ensure maximum benefit. We also incorporated a range of possible testing delays into a single number, the TAT. In reality, the delay could come from laboratories being overloaded with samples, issues with logistics or delays in contact tracing. Further, there are strategies that can increase the effectiveness of testing, by broadening the number of people required to quarantine when a case is identified. For example, in Victoria, Australia, rather than only identifying close contacts the Department of Health and Human Services has trialled quarantining the close contacts of a case, along with those individuals’ ‘second order’ close contacts (Mills & Clayton 2020). The impact of quarantining more people is currently unclear, but it does appear to be a strategy that further increases the impact of testing on overall transmission.

This work highlights how a testing strategy must complement general restrictions and that restrictions dictate the optimal priority ordering of tests. Testing and contact tracing capacity represent a finite (albeit expandable) resource, which should be targeted towards the most valuable cohorts. The average number of contacts for each person – and thus the potential reduction of transmission through contact tracing and quarantine – depends on what industries are open and what activities are allowed. The greater the spread potential, the greater the potential impact of testing and quarantining contacts, with the drawback of increased demands on the system. The optimal allocation of test efforts to reduce transmission also depends on community restrictions and the policy reaction once a case is identified. The benefit in transmission reduction from isolation will be comparatively larger if there are few community restrictions, and smaller if heavy restrictions, such as lockdowns, are already in place.

Metrics that track system performance must be chosen carefully, so that they support the objectives of a testing strategy. Performance measurement creates incentives that shape investment, but if the metrics are not well aligned with the objectives then performance can suffer. For example, reporting the percent positive and the number of tests conducted could encourage an increase in testing, which could be problematic if the true objective is to minimise transmission. Although reducing transmission is one of the five objectives of the European Union testing strategy (European Centre for Disease Prevention and Control 2020), the public data reports cases, number of tests and percent positive, but does not report metrics that relate to system efficiency (ECDC 2021).

Our results show that it is critical to have clear objectives to guide a testing strategy. While no one objective is ‘correct’ as approaches and aims vary between countries, having a clearly stated set of objectives and actions is important for developing effective management plans (Baker et al. 2020). Further, chosen objectives should be evaluable by reporting metrics. While developing an effective testing strategy is a challenging problem, focusing on quantifying the value of a test towards achieving the objectives is a practical approach for prioritising testing resources.

## Data Availability

All code used in this manuscript are available on github.

https://github.com/cmbaker00/optimal-test-allocation

## Code availability

The python code used to generate all results in this manuscript is available online https://github.com/cmbaker00/optimal-test-allocation.

## Acknowledgments

We would like to thank James McCaw for valuable discussions. We would also like to acknowledge funding from The Australian Government Department of Health, the NHMRC Centre of Research Excellence for Supporting Participatory Evidence generation to Control Transmissible diseases in our Region Using Modelling (SPECTRUM), The Melbourne Centre for Data Science (MCDS) and the Centre of Excellence for Biosecurity Risk Analysis (CEBRA). Jodie McVernon was supported by a NHMRC Principal Research Fellowship GNT1117140.

