## Supplementary information for "Optimal allocation of PCR tests to minimise disease transmission through contact tracing and quarantine"

**Optimal allocation of PCR tests to minimise transmission in an epidemic: supporting information**

#### *S1: Turnaround time and test volume*

We are not aware of any analyses that quantify the relationship between test volume and turnaround time. The show that a relationship exists and show that it's reasonable to assume that turnaround time increases with number of tests we use data from the UK testing system (Department of Health & Social Care 2020). As testing systems worldwide have been constantly changing, we would expect routine capacity to be also changing through time. Hence, we focus on a subset of the data, and we plot the data from the final 9 weeks of 2020 – the weeks ending in November and December. We use table 4 of the statistics "NHS Test and Trace statistics, 28 May to 30 December 2020: data tables" (Department of Health & Social Care 2020). We calculate the number of tests in each week by summing 'Number of test results received within 24 hours of taking a test', 'Number of test results received between 24 hours and 48 hours of taking a test', 'Number of test results received between 48 hours and 72 hours of taking a test', 'Number of test results received after 72 hours of taking a test', 'Number of test results communicated but unsuccessful' and 'Number of test results in progress not communicated yet'. The data does not provide the average turnaround time, instead breaking the data into four time windows. Instead of trying to estimate the average turnaround time, we show how the percentage of test results in each window changes as the number of tests increases, shifting away from sub-24 hours toward longer times.

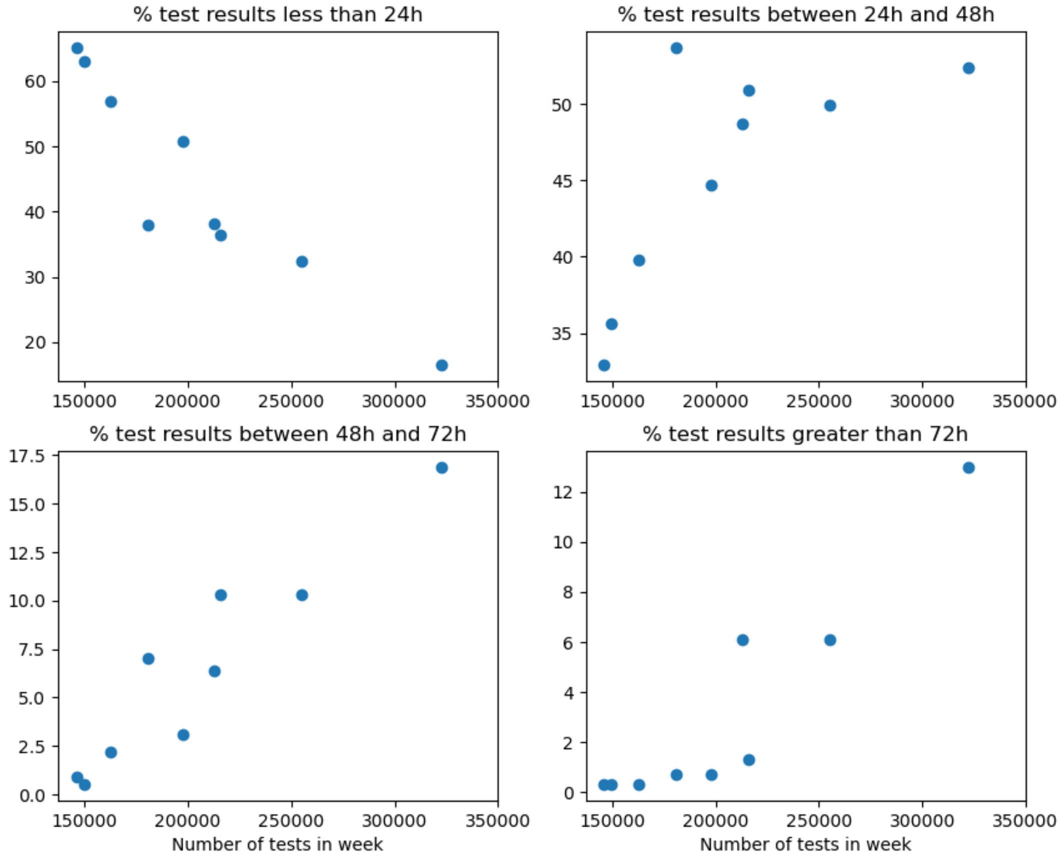


*Figure 1: NHS data from November and December 2020. The more tests conducted, the greater the delay between the test and the result.*

### *S2: Sensitivity to choice of function to model TAT*

In this section we explore the importance of the specified function to model TAT as the number of tests is varied. Throughout the manuscript we assume the TAT is constant below routine capacity, and increase quadratically above routine capacity, such that the TAT when the number of daily tests is 150% of routine capacity is 2 days. In the following figures we show results for onwards transmission using quadratic, linear and exponential functions to model the increase in TAT above routine capacity. Figure 2v and Figure 3 show the results for routine capacity of 2 tests per 1000, while Figure 4 and Figure 5 show the results with routine capacity of 4 tests per 1000. Across the three functions, there are some differences in the exact shape of the curves, but the important features of the results stay the same. In particular, the minimum transmission in each scenario is almost the same across each function.

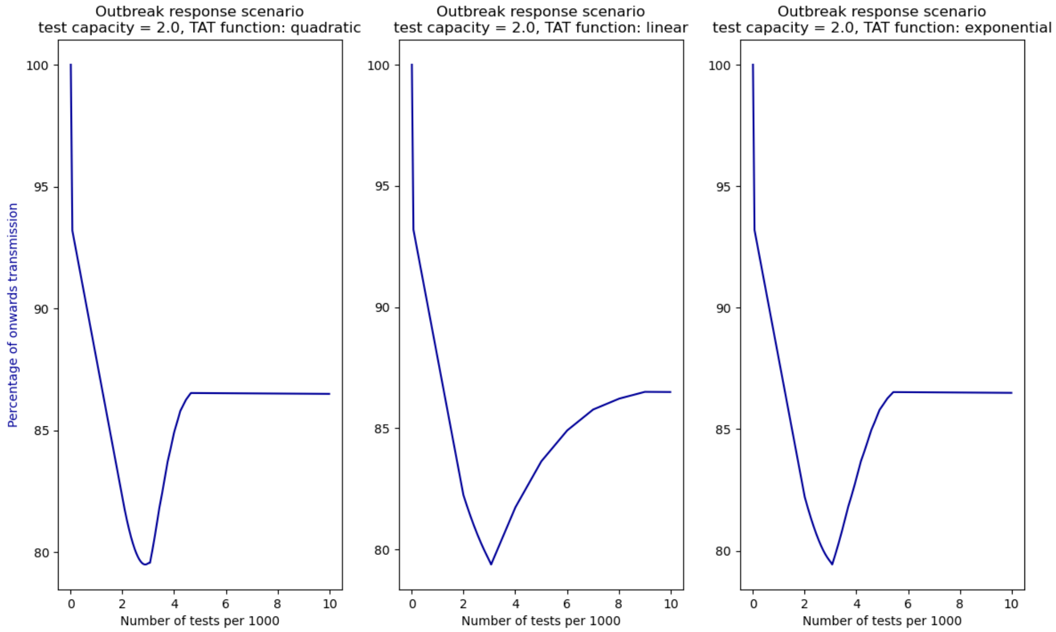


*Figure 2: Onwards transmission for the outbreak response scenario and routine capacity of 2 tests per 1000 per day, across three functions to model TAT as a function of test volume. The left plot shows the quadratic model, the middle plot is the linear function and the right plot is the exponential function.*

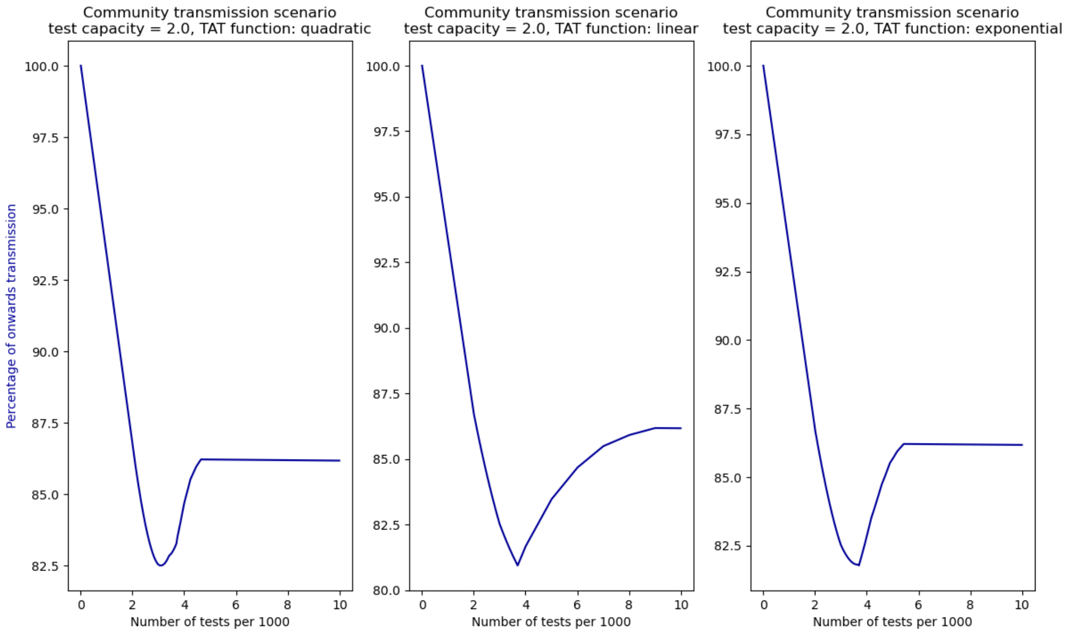


*Figure 3: Onwards transmission for the community transmission scenario and routine capacity of 2 tests per 1000 per day, across three functions to model TAT as a function of test volume. The left plot shows the quadratic model, the middle plot is the linear function and the right plot is the exponential function.*

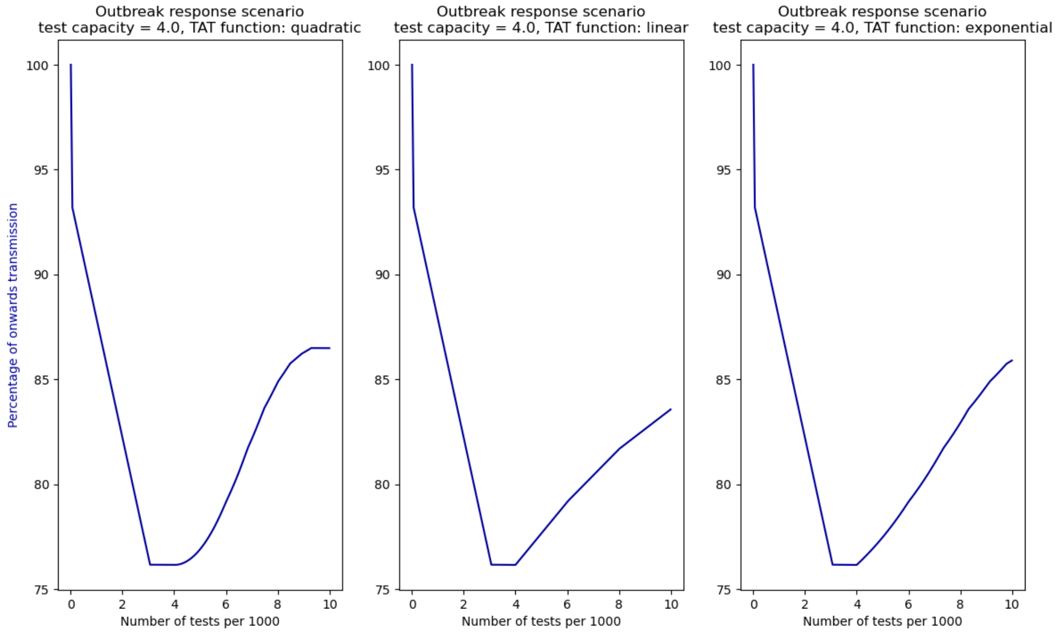

*Figure 4: Onwards transmission for the outbreak response scenario and routine capacity of 4 tests per 1000 per day, across three functions to model TAT as a function of test volume. The left plot shows the quadratic model, the middle plot is the linear function and the right plot is the exponential function.*

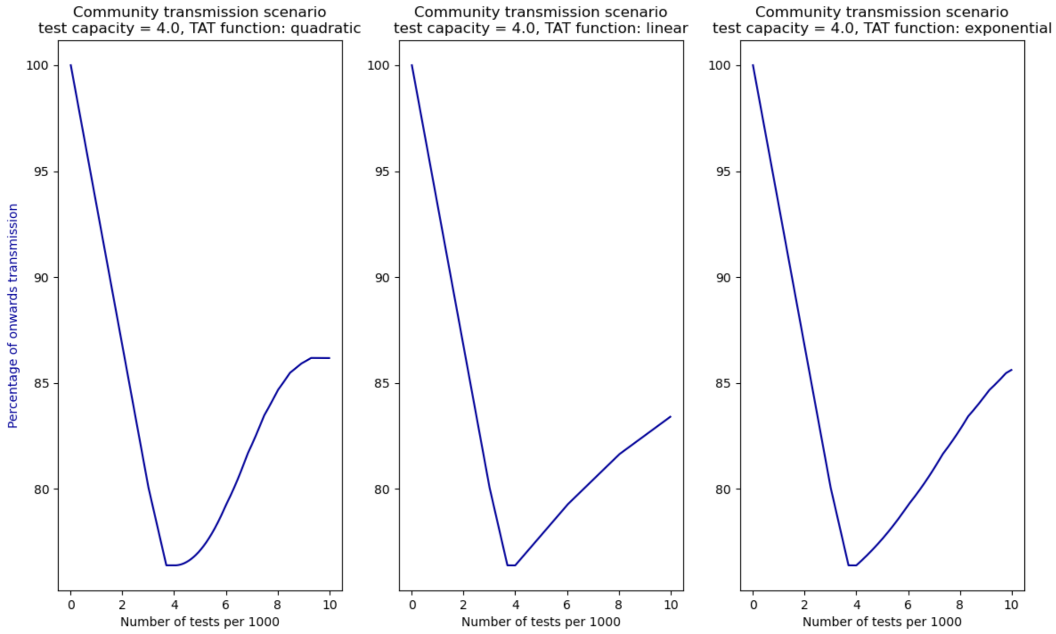


*Figure 5: Onwards transmission for the community transmission scenario and routine capacity of 4 tests per 1000 per day, across three functions to model TAT as a function of test volume. The left plot shows the quadratic model, the middle plot is the linear function and the right plot is the exponential function.*

#### *S3: Symptomatic presenting proportion*

This section provides addition plots, showing the reduction in transmission and the percentage of infections identified across different proportions of symptomatic presentations. The proportion of people with COVID-19 symptoms that actually get tested is highly uncertain and likely varies significantly globally. The 12 plots included here show our results for the presenting proportion being 25%, 50% and 75%, across our two epidemic scenarios and two testing capacity values.

There are two important points raised in these figures. The first is that for the outbreak response scenario, the percentage of infections identified is very low, ranging from just over 2% (Figure 6 and Figure 12) to just over 6% (Figure 8 and Figure 14). The driver of the low case identification percentage is that there are very few cases in even close contacts and symptomatic people, and there is not sufficient testing capacity to test a large proportion of the asymptomatic population. Secondly, all plots, except one, show the same qualitative behaviour, where it is best to test at, or slightly above, routine capacity. The only exception is for the community transmission scenario when test capacity is low and the presenting proportion is high (Figure 11). In this scenario, it is optimal in the model to test as many people as possible. The mechanism is that the model suggests using testing to get as many people as possible to isolate, and getting every close contact and symptomatic person to isolate is more beneficial than maximising contact tracing efficiency.

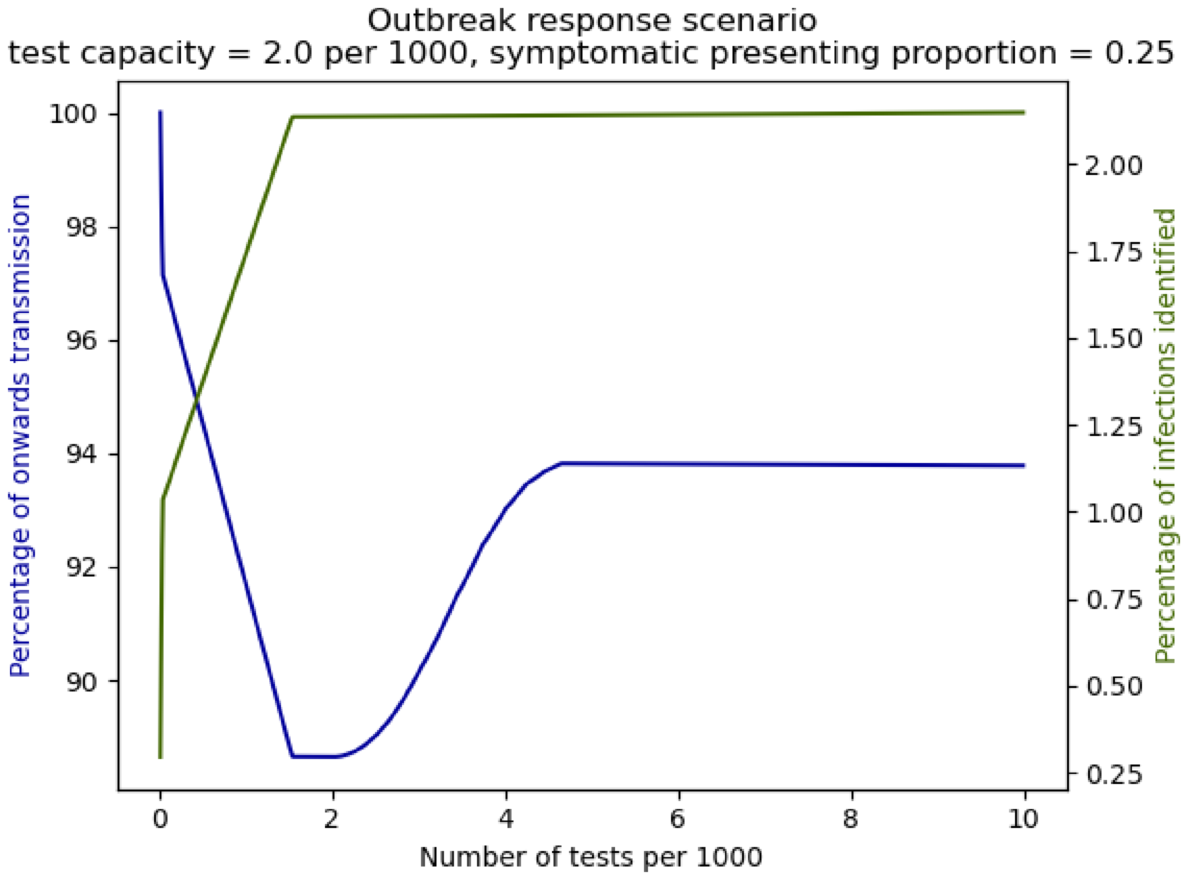


*Figure 6: Reduction in transmission (solid blue line) for the outbreak response scenario with test capacity of 2 per 1000 per day and 25% of symptomatics available for testing. The solid green line shows the percentage of total infections that are identified as cases.*

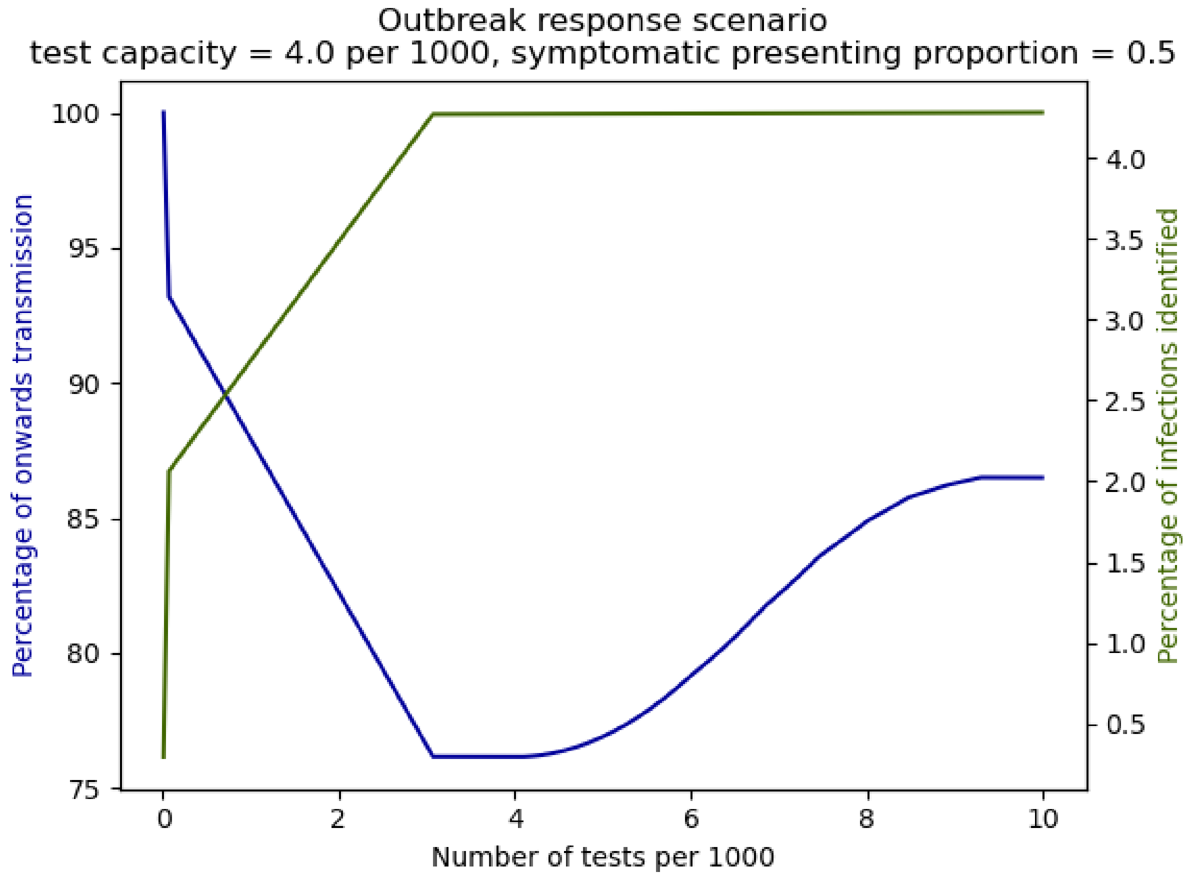


*Figure 7: Reduction in transmission (solid blue line) for the outbreak response scenario with test capacity of 2 per 1000 per day and 50% of symptomatics available for testing. The solid green line shows the percentage of total infections that are identified as cases.*

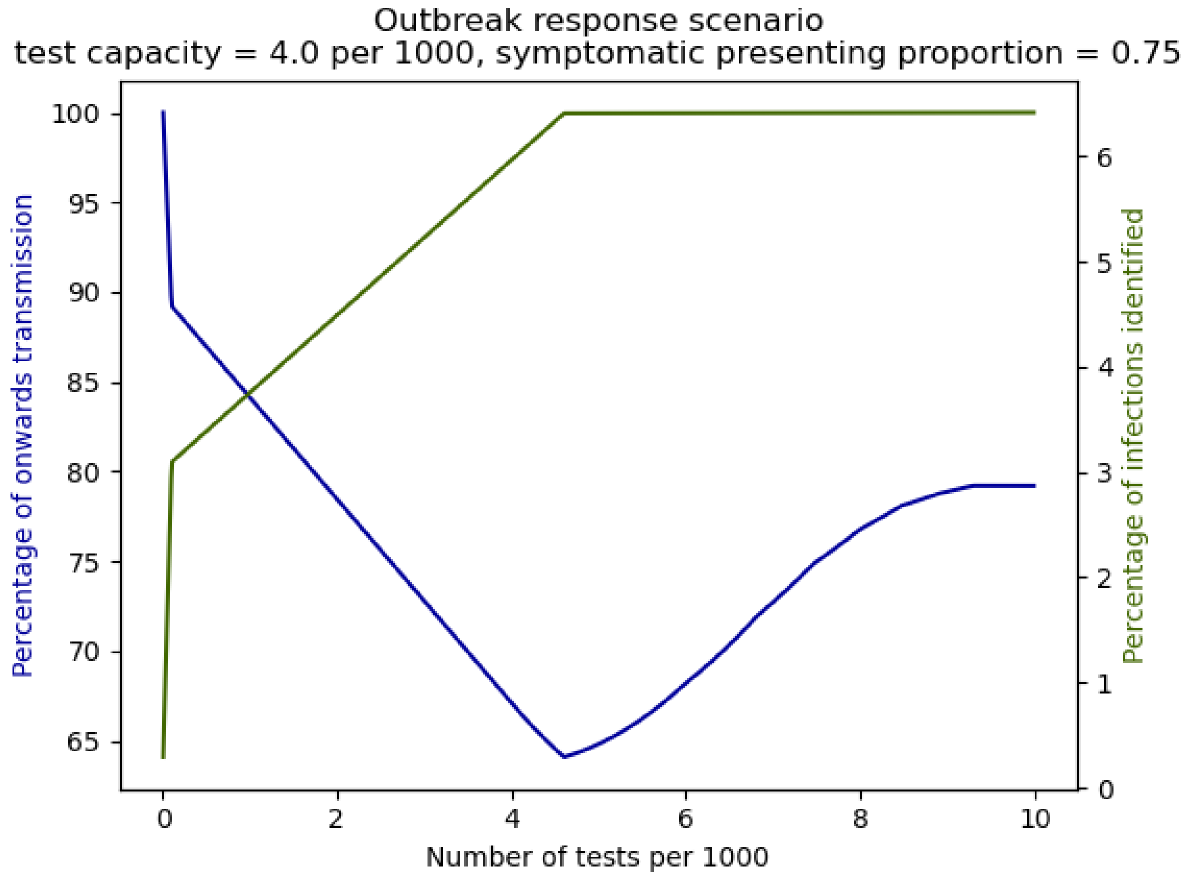


*Figure 8: Reduction in transmission (solid blue line) for the outbreak response scenario with test capacity of 2 per 1000 per day and 75% of symptomatics available for testing. The solid green line shows the percentage of total infections that are identified as cases.*

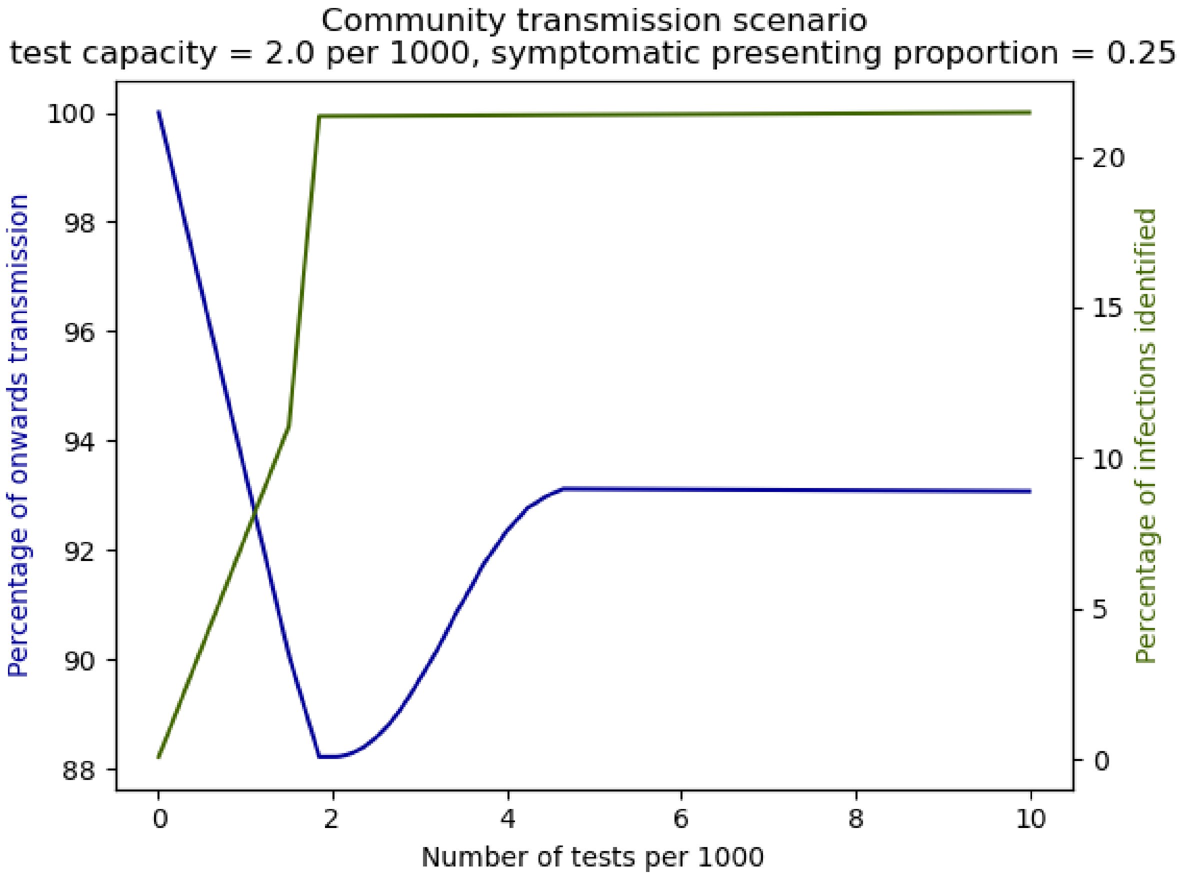


*Figure 9: Reduction in transmission (solid blue line) for the community transmission scenario with test capacity of 2 per 1000 per day and 25% of symptomatics available for testing. The solid green line shows the percentage of total infections that are identified as cases.*

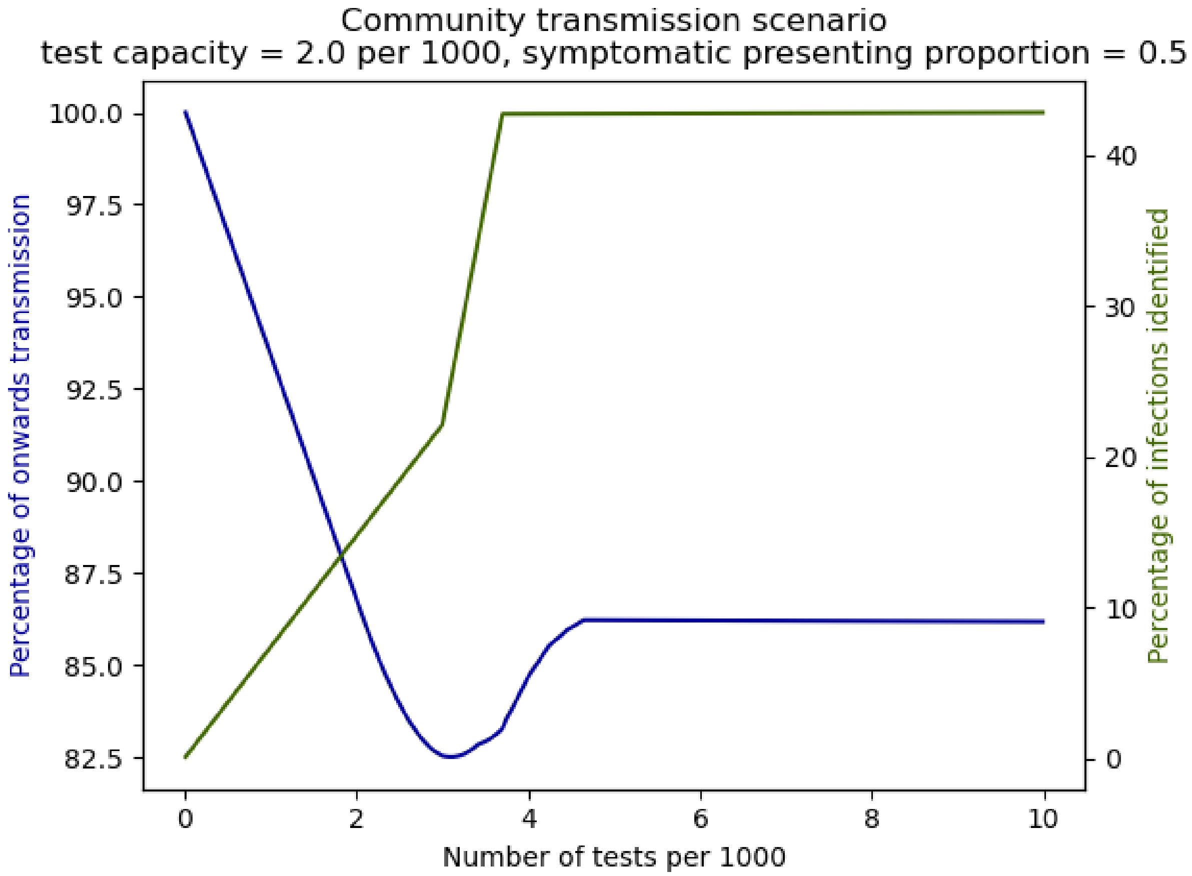


*Figure 10: Reduction in transmission (solid blue line) for the community transmission scenario with test capacity of 2 per 1000 per day and 50% of symptomatics available for testing. The solid green line shows the percentage of total infections that are identified as cases.*

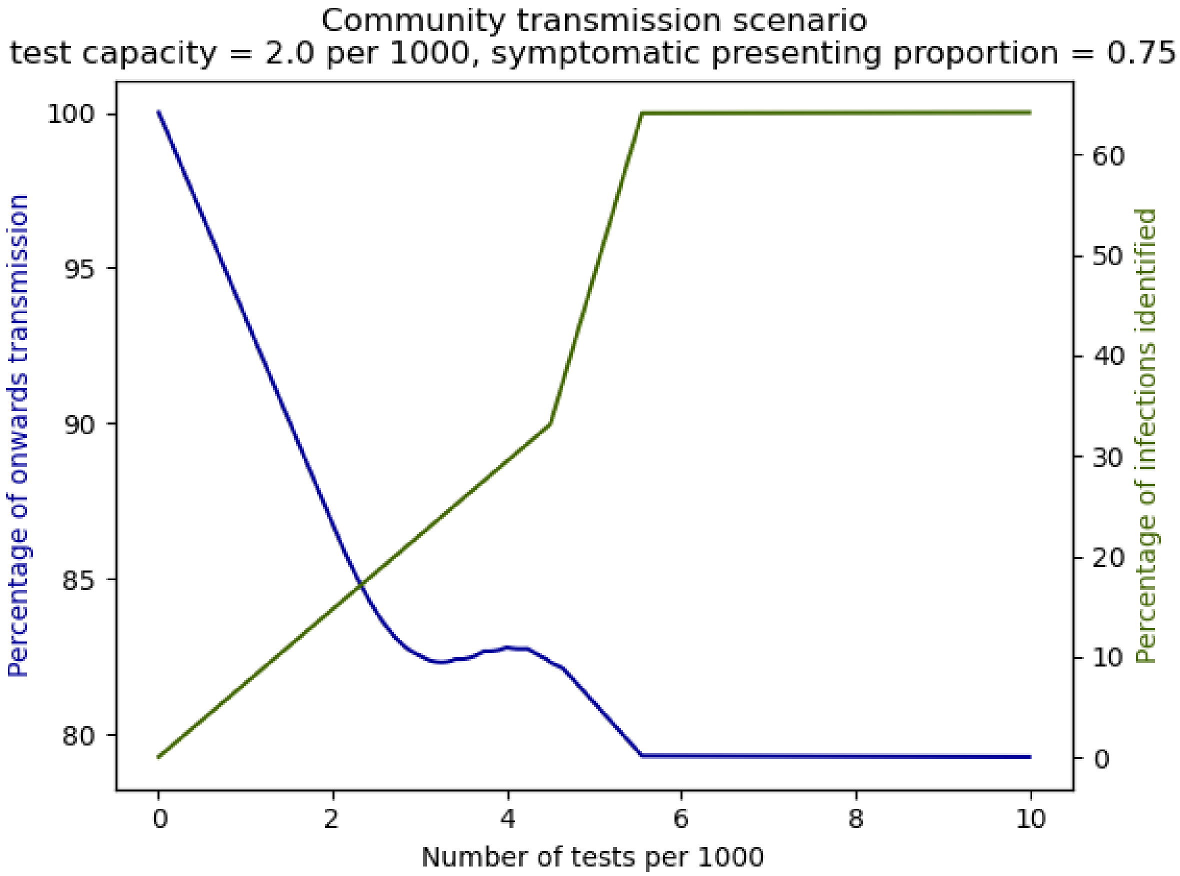


*Figure 11: Reduction in transmission (solid blue line) for the community transmission scenario with test capacity of 2 per 1000 per day and 75% of symptomatics available for testing. The solid green line shows the percentage of total infections that are identified as cases.*

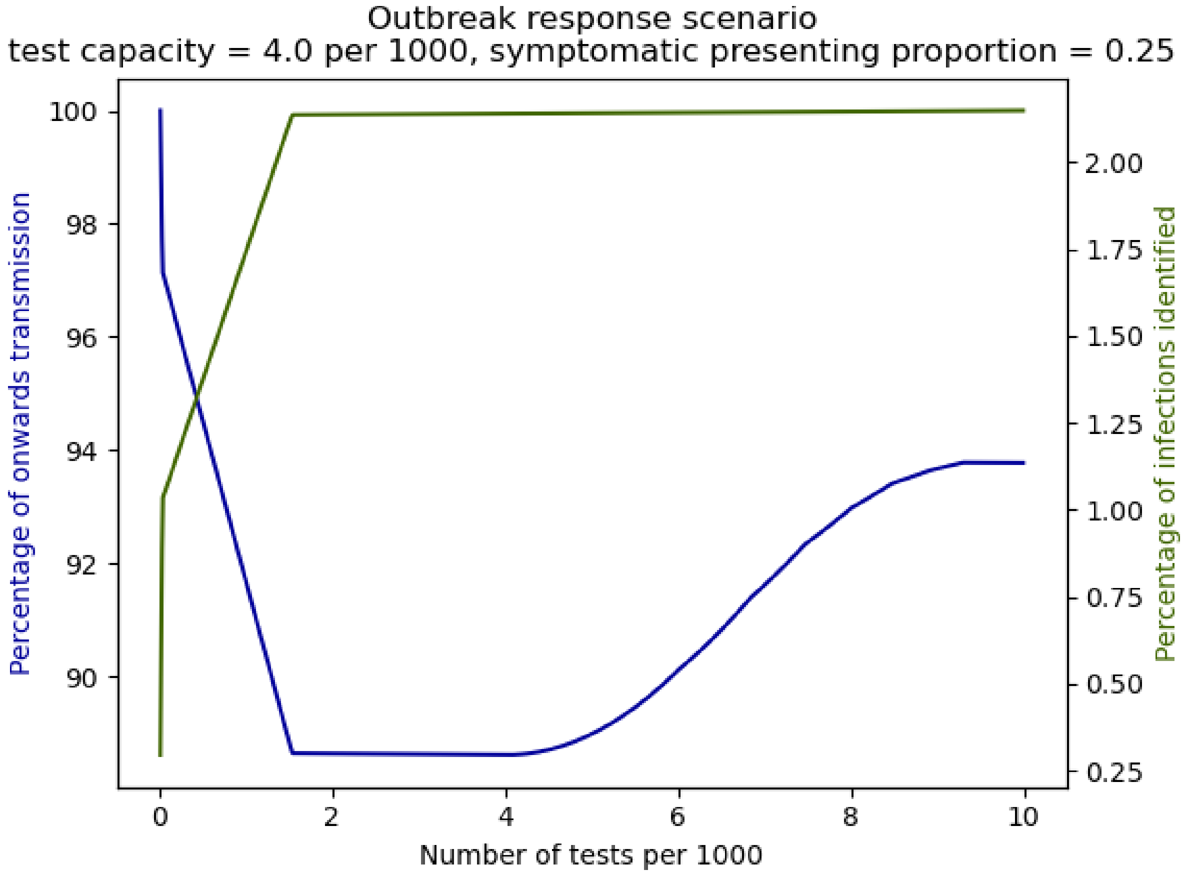


*Figure 12: Reduction in transmission (solid blue line) for the outbreak response scenario with test capacity of 4 per 1000 per day and 25% of symptomatics available for testing. The solid green line shows the percentage of total infections that are identified as cases.*

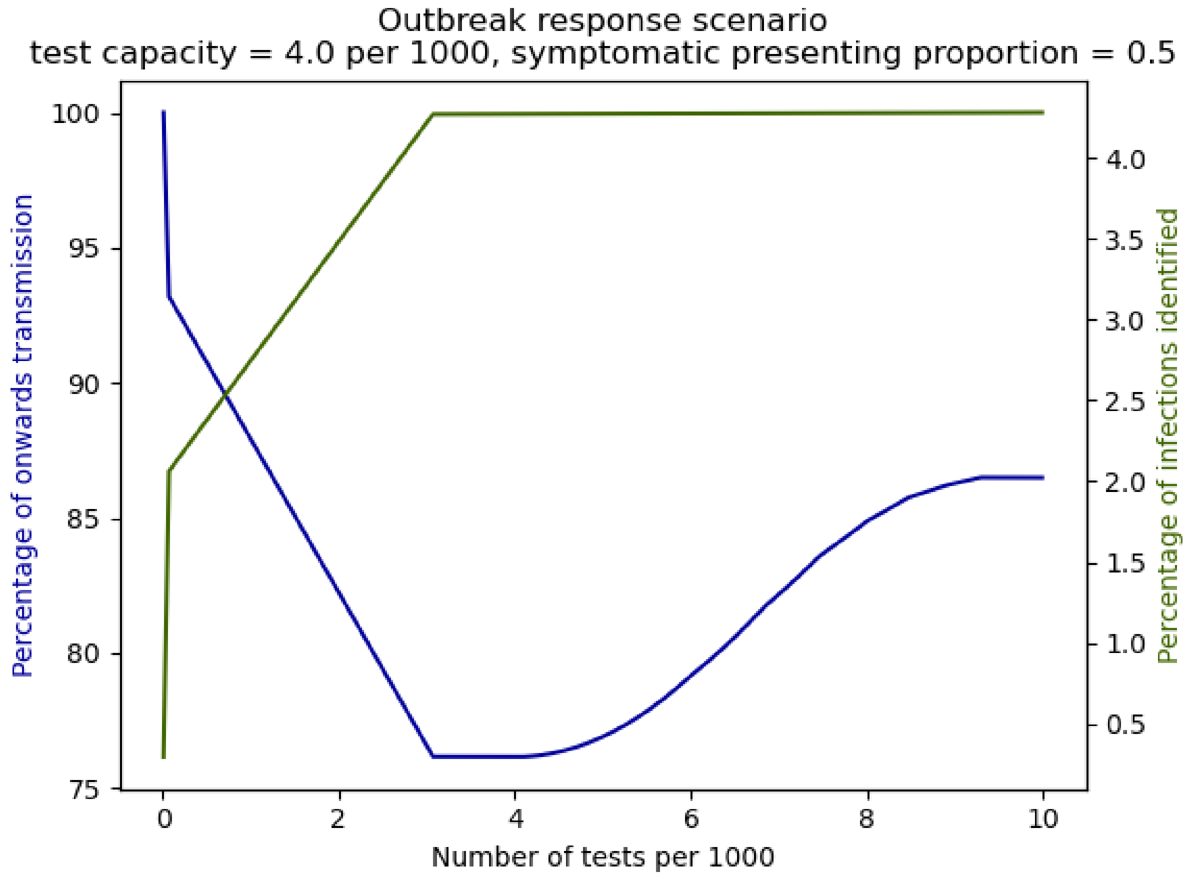


*Figure 13: Reduction in transmission (solid blue line) for the outbreak response scenario with test capacity of 4 per 1000 per day and 50% of symptomatics available for testing. The solid green line shows the percentage of total infections that are identified as cases.*

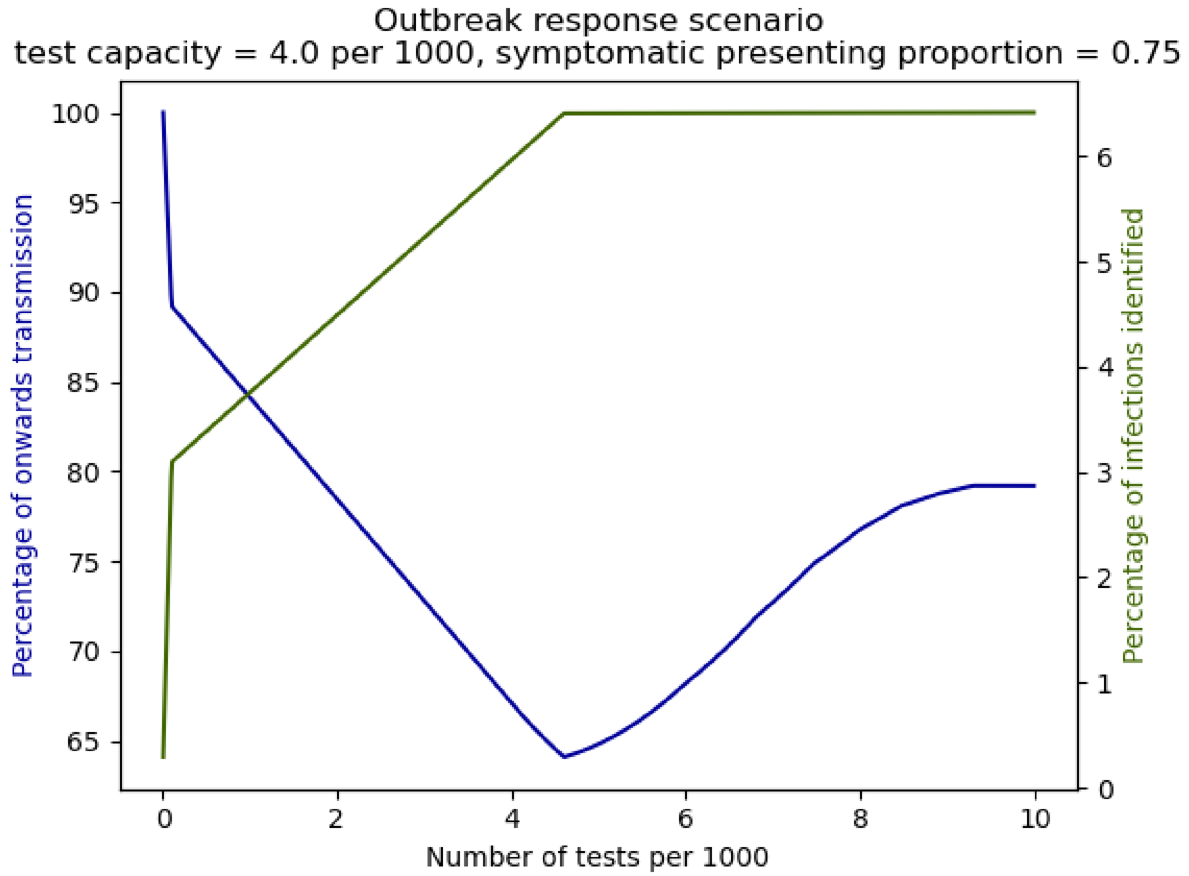


*Figure 14: Reduction in transmission (solid blue line) for the outbreak response scenario with test capacity of 4 per 1000 per day and 75% of symptomatics available for testing. The solid green line shows the percentage of total infections that are identified as cases.*

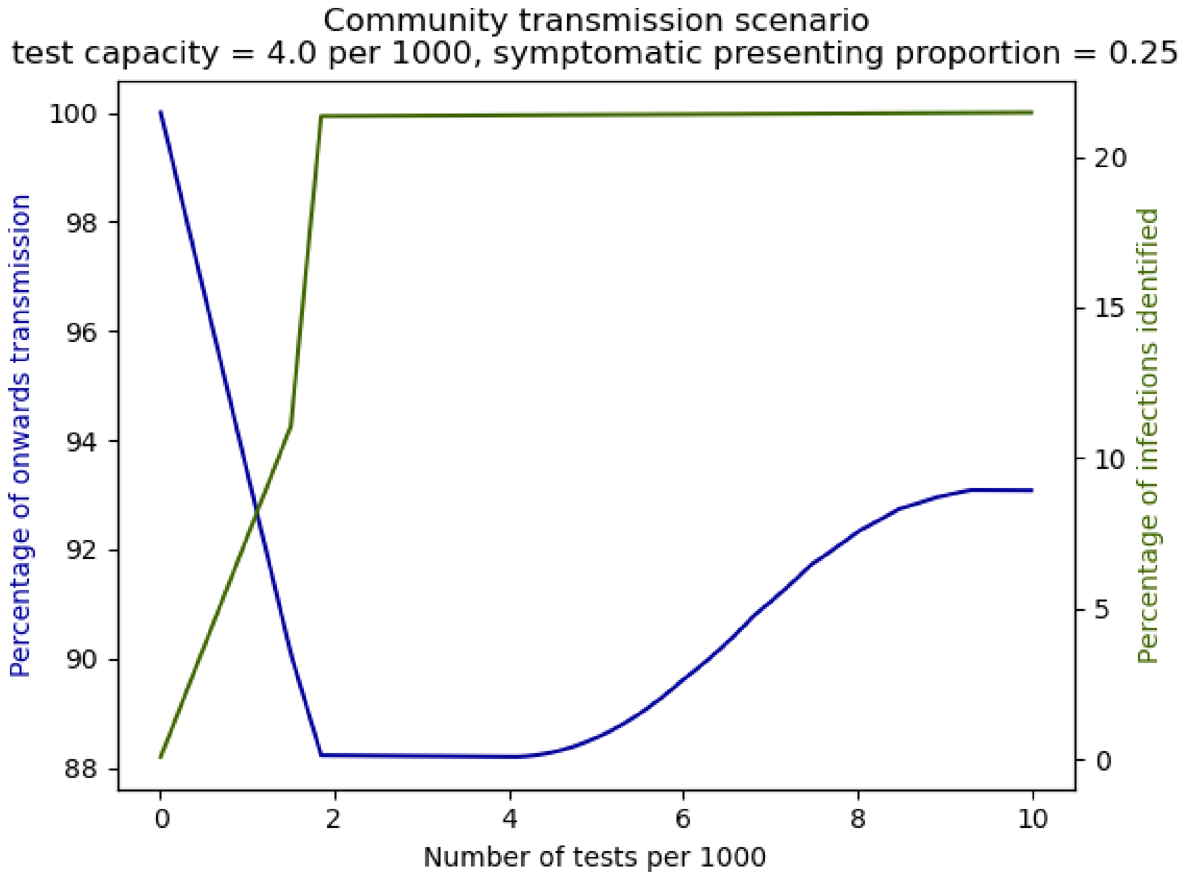


*Figure 15: Reduction in transmission (solid blue line) for the community transmission scenario with test capacity of 4 per 1000 per day and 25% of symptomatics available for testing. The solid green line shows the percentage of total infections that are identified as cases.*

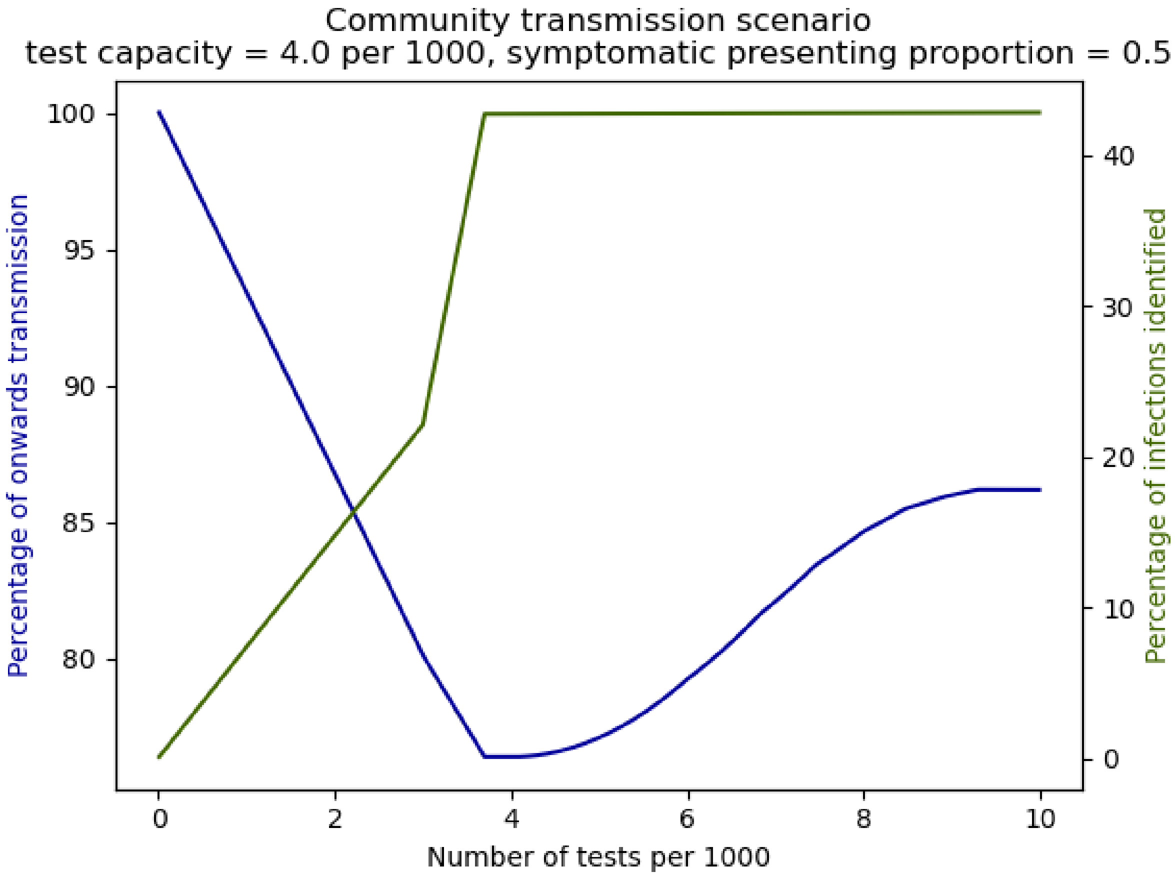


*Figure 16: Reduction in transmission (solid blue line) for the community transmission scenario with test capacity of 4 per 1000 per day and 50% of symptomatics available for testing. The solid green line shows the percentage of total infections that are identified as cases.*

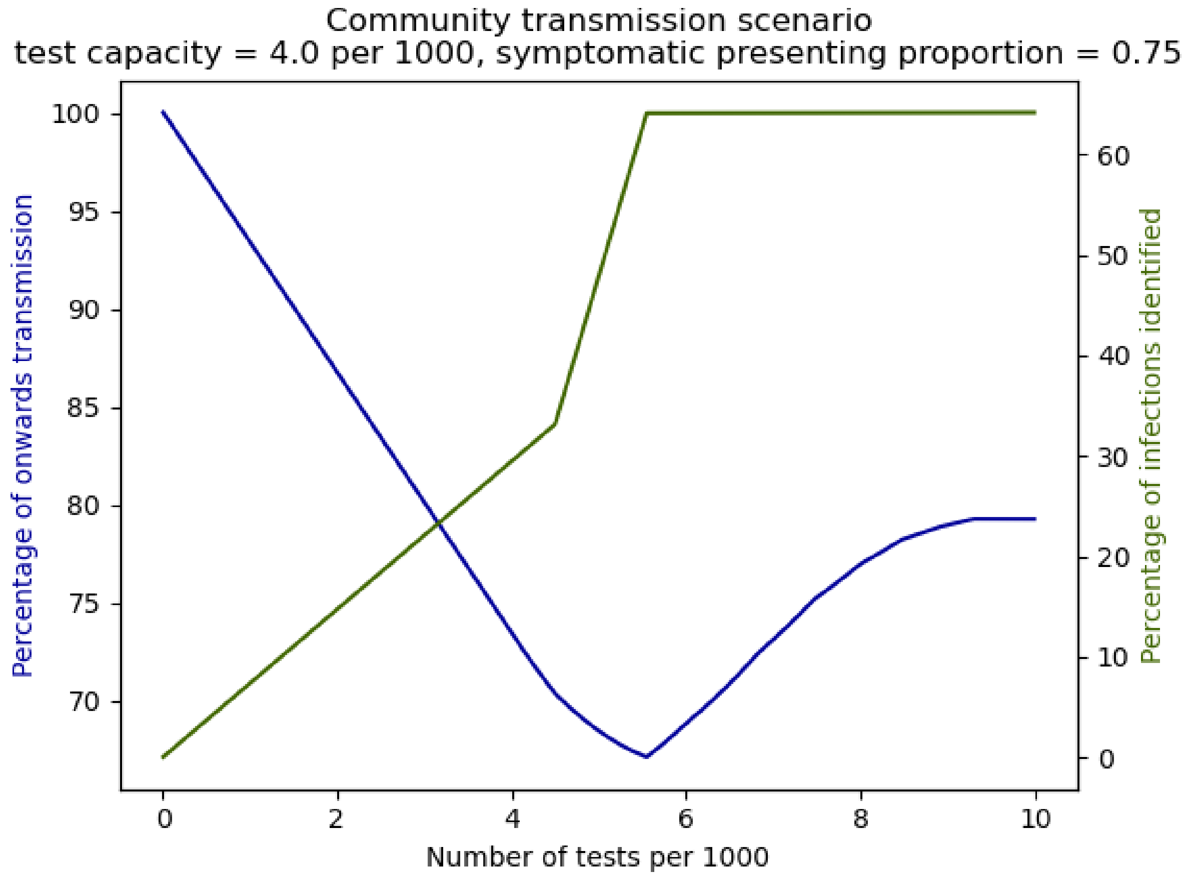


*Figure 17: Reduction in transmission (solid blue line) for the community transmission scenario with test capacity of 4 per 1000 per day and 75% of symptomatics available for testing. The solid green line shows the percentage of total infections that are identified as cases.*

*S4: Sensitivity to onwards transmission, population distribution and infection probability*

In this section we provide plots to show how the population onwards transmission depends on: each groups' onwards transmission (Figure 18); the proportion of the population in each group (close contact, symptomatic, asymptomatic) (Figure 19), and the probability of a person being infected (Figure 20) affects the results. We vary each of these parameters one at a time, using the community transmission scenario with 4 tests per 1000 per day as the test capacity. For each of the plots we sample from a uniform distribution with minimum and maximum values 20% below or above their default values respectively. We do 100 simulations for each plot and the shaded area includes the 5$^{th}$ to 95$^{th}$ percentile of parameter values.

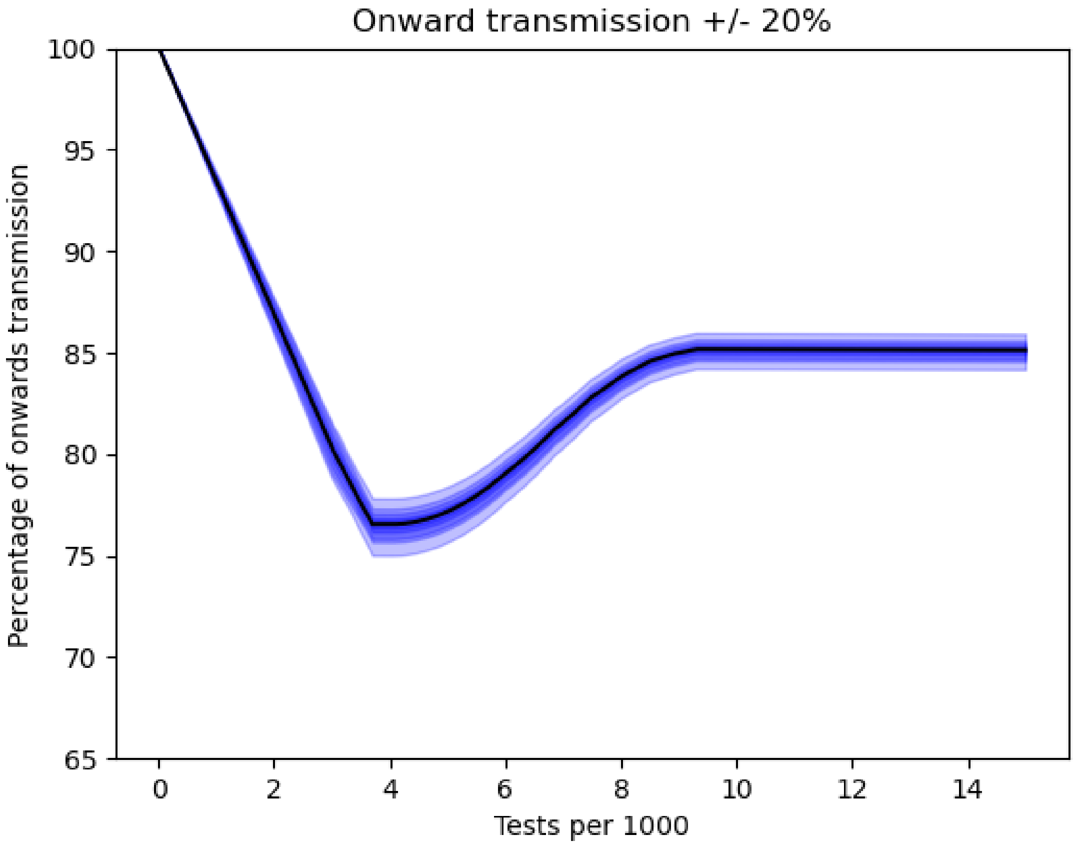


*Figure 18: The percentage of onwards transmission as the amount of onwards transmission is varied for each of the groups.*

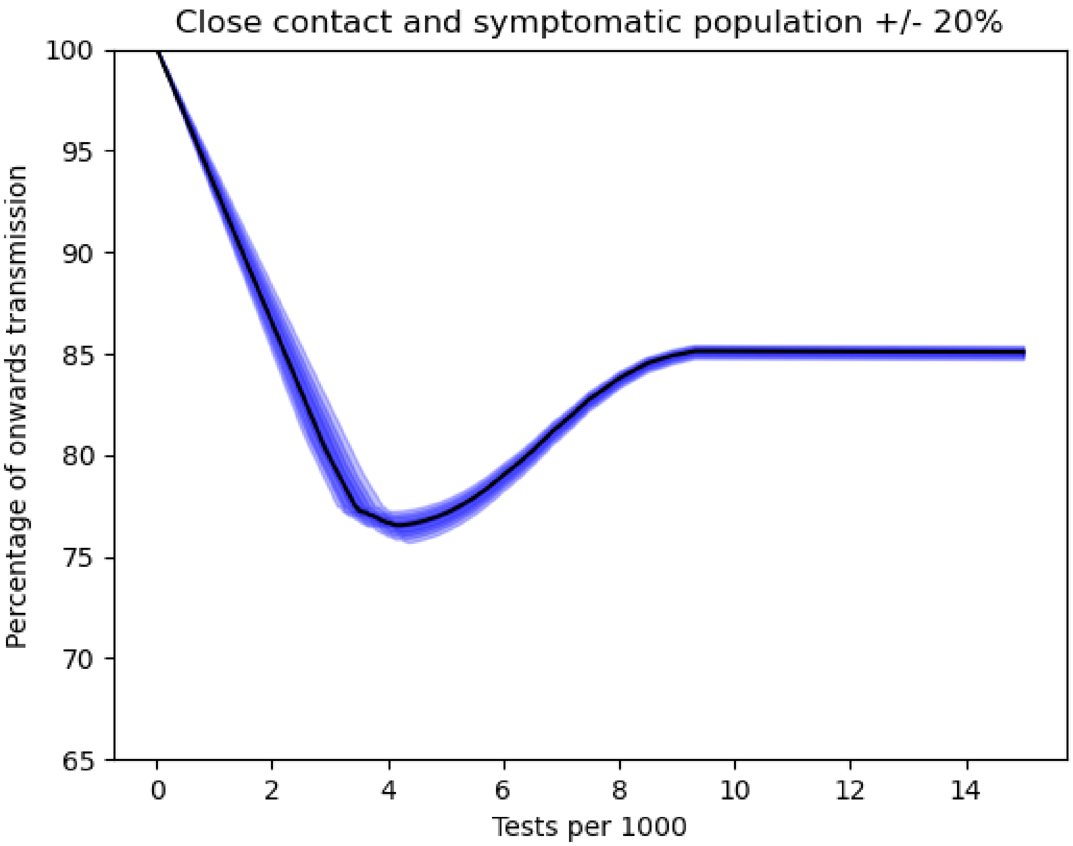


*Figure 19: The percentage of onwards transmission as the amount of people in the close contact and symptomatic groups is varied.*

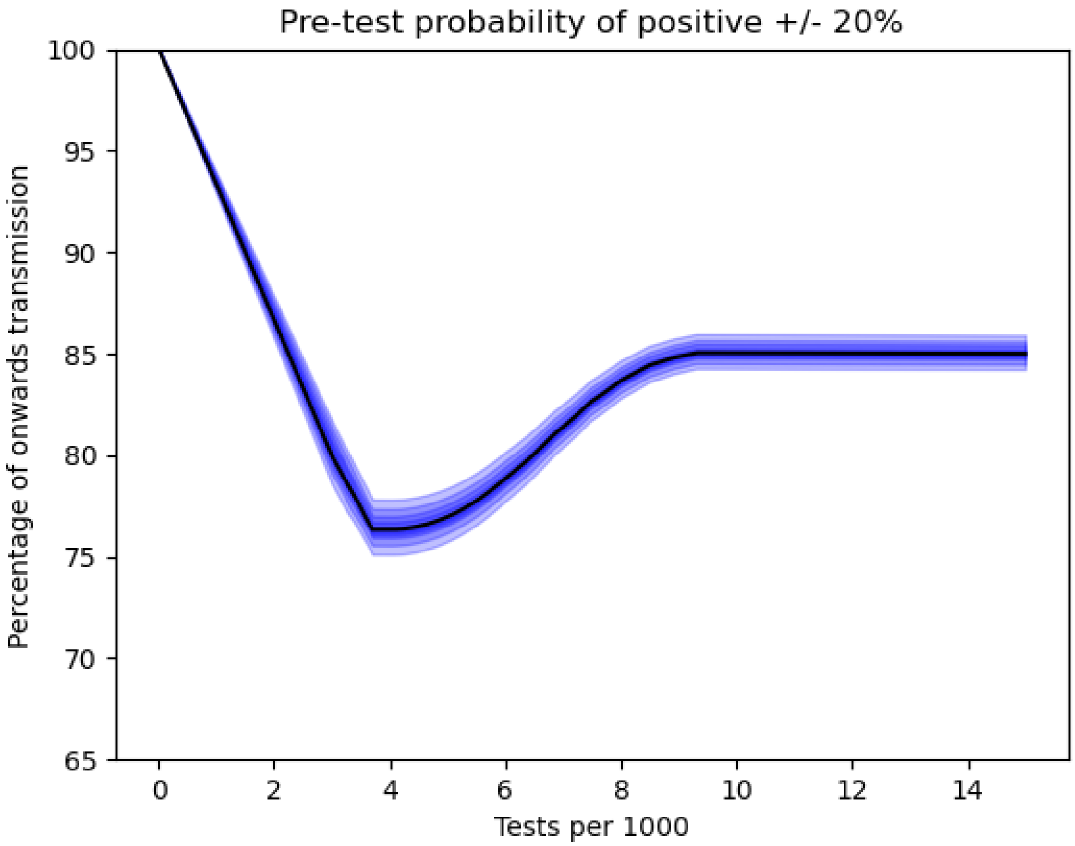


*Figure 20: The percentage of onwards transmission as the probability of testing positive in each group is varied.*

***References***

Department of Health & Social Care. (2020). *NHS Test and Trace (England) statistics: 24 December to 30 December*.
